# Deep learning classification of MRI differentiates brain changes in genetic and idiopathic Parkinson’s disease

**DOI:** 10.1101/2024.11.21.24317644

**Authors:** Megan Courtman, Mark Thurston, Hongrui Wang, Sube Banerjee, Adam Streeter, Lucy McGavin, Stephen Hall, Lingfen Sun, Emmanuel Ifeachor, Stephen Mullin

**Author notes:** Correspondence to: Megan Courtman, Room N6, Phase I building, Plymouth Science Park, 1 Davy Rd, Plymouth, PL6 8BX. **Funding sources:** This work was supported by the Engineering and Physical Sciences Research Council under Grant EP/T518153/1 and the National Institute for Health and Care Research under Grant PEN/006/005/A.

## Abstract

**Background:** The prospect of neuroprotective treatments for Parkinson’s disease highlights the need for early diagnostic tests. Specialised MRI sequences suggest changes related to Parkinson’s disease may be detectable.

**Objectives:** We used the Parkinson’s Progression Markers Initiative dataset to investigate whether deep learning can detect early brain MRI changes of idiopathic and GBA/LRRK2 prodromal Parkinson’s disease.

**Methods:** Pairs of matched cohorts were used to train convolutional neural networks to classify T2 axial images. Explainability methods were used to visualise drivers of model predictions.

**Results:** Models built to distinguish between idiopathic Parkinson’s disease scans (*n*=504) and matched controls exhibited good classification performance for scans taken more than four years after diagnosis, with a Receiver Operating Characteristic area-under-the-curve of 0.89 (*n*=98). Model performance deteriorated as time since diagnosis reduced. Models built to distinguish non-manifesting carriers of *LRRK2* (area-under-the-curve 0.92, 90% accuracy, *n*=115) and *GBA* (area-under-the-curve 0.94, 92% accuracy, *n*=109) from controls exhibited excellent classification performance. All models demonstrated foci of interest in cerebrospinal fluid spaces surrounding the brainstem. Models using *GBA* scans also identified foci of interest in occipital lobes.

**Conclusions:** Deep learning models appear able to reproducibly detect changes in the brains of those with established but not early Parkinson’s disease. Conversely changes in at risk genetic cohorts are detectable at all stages, including in those who have not developed Parkinson’s disease. This implies a distinct pathological process ongoing within the brains of carriers of Parkinson’s disease genetic risk factors compared to those with sporadic Parkinson’s disease.

## Introduction

The prospect of disease-modifying treatments for Parkinson’s disease highlights the need for early diagnostic tests. A number of neuroprotective compounds have yielded promising results in phase II clinical trials.^1^ These have the potential to prevent the death of dopaminergic neurons and avoid the most debilitating movement related (motor) features of Parkinson’s disease. To be most effective, they would need to be administered as early as possible, ideally in the prodromal period of Parkinson’s disease, which can precede the onset of motor symptoms by a decade or more.^2^ During this time patients may experience a variety of non-motor features in advance of the diagnostic motor features of Parkinson’s disease.^3^ Studies suggest that 30-50% of dopaminergic neurons within the basal ganglia die before these motor features become clinically significant.^4–6^

Much research into the prodromal phase of Parkinson’s disease has focussed on those with the genetic risk factors glucocerebrosidase (*GBA*) and Leucine-rich repeat kinase 2 (*LRRK2*). These risk factors are distinct from ‘monogenic’ forms of Parkinson’s disease, which are rare, transmit in a Mendelian manner with near 100% penetrance and can manifest as early as the third decade. Conversely *GBA and LRRK2* have a penetrance of 8-10% and 28-74% respectively.^7,8^ *LRRK2* Parkinson’s disease manifests at an average age of 59.4,^9^ and is thought to progress more slowly, often with a milder tremor predominant phenotype. *GBA* manifests at an average age of 55.8,^9^ is associated with more cognitive/neuropsychiatric symptoms and tends to progress more rapidly.^10^ Disease phenotype in *GBA* appears to be variant dependent. Based on a classification of symptoms documented in cases of the autosomal recessive lysosomal storage disorder Gaucher disease (caused by *GBA* variants in a biallelic state), *GBA* risk variants can be classified as ‘severe’, ‘mild’ and non-Gaucher causing Parkinson’s disease risk variants. It has been shown that ‘severe’ *GBA* entail a higher disease risk with more rapid progression of Parkinson’s disease in those who develop it.^10^

Parkinson’s disease remains a clinical diagnosis. MRI is not in widespread use as a diagnostic tool in Parkinson’s disease, although promising evidence exists of early changes detectable on specialised MRI sequences.^11^ Analysis of MRI brain scans undertaken for other purposes might provide a cost-effective means of identifying some with early Parkinson’s disease, particularly if such changes were present in prodromal cases. There is evidence to suggest that this might be feasible. Prodromal brain changes have been observed in other imaging modalities. Dopamine active transporter (DAT) scans carried out in subjects with hyposmia, REM sleep behaviour disorder (RBD) and non-Parkinson’s disease manifesting *LRRK2* variant carriers have demonstrated abnormal dopamine uptake compared to controls.^12^

A subset of machine learning known as deep learning may be able to detect patterns in scans imperceptible by the human visual system. Machine learning describes algorithms that improve themselves without explicit instruction through exposure to data. There are different types of machine learning, many of which require manual feature selection. Deep learning describes the use of layered neural networks to build representations of complicated concepts out of simpler concepts. A distinctive aspect of deep learning is that, unlike traditional machine learning, it does not require manual feature selection or engineering, so it can learn more abstract representations that might not have been anticipated by human practitioners. In diagnostic imaging, deep learning has shown great promise in the last decade, being able to match and in some cases outperform human medical practitioners for routine radiological reporting.^13^ Traditional pre-defined feature systems have not generally met the stringent performance requirements for clinical utility,^14^ but deep learning methods have achieved higher performances, allowing the deployment of artificial intelligence (AI) based applications in a number of defined clinical contexts.

Previous studies have used the Parkinson’s Progression Markers Initiative (PPMI) to investigate whether deep learning might be used to distinguish between Parkinson’s disease and control MRI scans.^15^ Such studies have reported accuracies as high as 100%^16^. However, a lack of explainability as to the regions of interest in these studies has led to major sources of confounding being overlooked. For example, in a substantial proportion of studies, serial scans from the same patient were included in both training and test datasets, creating a source of data leakage.^17^ There has also been little focus on the PPMI genetic cohorts.

In this report, we describe the development of deep learning models in both idiopathic Parkinson’s disease and *GBA/LRRK2* populations with and without Parkinson’s disease, designed to allow differentiation from age matched control scans. These models control for potential sources of data leakage, and leverage state of the art explainable artificial intelligence techniques to understand the predictions made, producing models with a greater level of sophistication and assurance.

## Materials and methods

### Study design and participants

This study uses data from the PPMI cohort.^18^ PPMI is an international observational study recruiting patients through outpatient neurology practices at academic centres in Austria, Canada, France, Germany, Greece, Israel, Italy, the Netherlands, Norway, Spain, the UK, and the USA, with the goal of identifying clinical and biological markers of disease heterogeneity and progression in Parkinson’s disease. The PPMI study is registered with ClinicalTrials.gov (number NCT01141023). Detailed information about inclusion criteria, informed consent, demographic data, and study design can be found on the PPMI website.

Participants in this study were included in one of four cohorts: idiopathic Parkinson’s disease (non-carriers of genetic variants associated with Parkinson’s disease), healthy controls, Parkinson’s disease manifesting carriers of *GBA* and *LRRK2* risk variants, and non-Parkinson’s disease manifesting carriers of *GBA* and *LRRK2* risk variants. The diagnosis for each group was made by site investigators who are movement disorder specialists and confirmed by a central consensus committee review. The PPMI study was approved by the institutional review board at each site, and participants provided written informed consent.

### Data selection

All available MRI studies (*n*=5988) were downloaded from the PPMI website on 25 November 2020, along with demographic and clinical data, including genetic status and date of Parkinson’s disease diagnosis. From this full MRI dataset, all T2-weighted axial scans were identified automatically using MRI parameters (Echo Time, Repetition Time) contained within the DICOM tags.

### Data organisation

The data were grouped into pairs of cohorts for the constructing of binary classification models. Cohorts to be compared were matched by age and sex. Ten-fold cross-validation was used to develop and assess models, with the data divided into 90% training data and 10% validation data in each fold. As many subjects have contributed more than one scan to the dataset, scans were grouped by subject before being divided so that the same subject never appeared in both the training and the test data.

### Image preprocessing

Scans were registered to a standard MNI template and skull-stripped using the FMRIB Software Library.^19^ All further preprocessing was carried out using the Python programming language. Volumes were cropped to the outermost dimensions of the brain and resized to 32×32×16 voxels. Voxel values were scaled between zero and one.

### Neural network architecture

Python-based deep neural networks were built with Keras using the TensorFlow backend.^20^ Graphics processing unit hardware acceleration was used for neural network training.

For each pair of cohorts, a three-dimensional convolutional neural network was trained from scratch, due to a lack of available pre-trained three-dimensional classification networks. To approximate the optimal network structure for these data, different hyperparameter configurations were trialled in the early stages. These hyperparameters were tuned following curve analysis at each iteration. Once no further reductions in the validation loss could be achieved, the hyperparameter configuration was finalised, and this architecture was used for all models (Supplementary Fig. 1).

### Model training

The models were trained for a maximum of 10000 epochs using stochastic gradient descent with the Adam optimisation algorithm.^21^ The binary cross-entropy loss function was utilised. Images were augmented with horizontal flip. Other augmentation methods were trialled but did not result in any further increase in performance. Early stopping with a patience of 500 was utilised. The models achieving the lowest loss on the validation sets during training were saved using checkpoints. A classification threshold was then chosen for the models which best balanced accuracy, sensitivity, and specificity.

### Explainability

SHapley Additive exPlanations (SHAP) were used to explain the models’ predictions. SHAP uses the game theory concept of Shapley values to calculate the contribution of a factor to a machine learning model output.^22^ In this case, SHAP was used to calculate and visualise the contribution of individual pixels to the deep learning model’s prediction. To visualise areas of interest, the Shapley values were used to generate explainable heatmaps for samples of individual cases, as well as average heatmaps for each comparison. All heatmaps were scaled to the same range of values, and average heatmaps were overlaid on a scan from a 64-year-old control subject (the average age of included subjects).

## Results

### Idiopathic Parkinson’s disease

In this analysis, 504 scans from 193 subjects with idiopathic Parkinson’s disease were used. All subjects had undergone genetic testing for *LRRK2*, *GBA* or *SNCA* mutations with no pathological or PD risk factor variants found. To investigate whether model performance is affected by disease progression, these scans were stratified by time since diagnosis: those acquired more than four years after diagnosis (*n*=98), those acquired two to four years after diagnosis (*n*=133), those acquired one to two years after diagnosis (*n*=122), and those acquired less than a year after diagnosis (*n*=151). Each of these cohorts was matched on age and sex with healthy control scans in a ratio of 1:1. Demographic data for these cohorts are shown in Table 1. Classification thresholds were chosen to maximise accuracy and balance sensitivity and specificity (Supplementary Fig. 5).

**Table 1:** Patient data for compared cohorts. PD = Parkinson’s disease. IPD = idiopathic PD. *LRRK2* PD = *LRRK2* PD manifesting carriers. *LRRK2* nPD = *LRRK2* non PD manifesting carriers. *GBA* PD = *GBA* PD manifesting carriers. *GBA* nPD = *GBA* non PD manifesting carriers. GC *GBA* nPD = Gaucher causing *GBA* variants non manifesting carriers.

| Compared cohorts | Cohort size | Median age (interquartile range) | % male | Median symptom duration in months (interquartile range) | % Dementia | % Mild cognitive impairment |
| --- | --- | --- | --- | --- | --- | --- |
| IPD < 1 year | 151 | 63 (55-69) | 69 | 14 (10-25) | 0 | 16 |
| Matched controls | 151 | 62 (56-69) | 69 | - | 0 | 1 |
| IPD 1-2 years | 122 | 63 (54-69) | 69 | 28 (23-40) | 0 | 19 |
| Matched controls | 122 | 62 (54-69) | 69 | - | 0 | 2 |
| IPD 2-4 years | 133 | 65 (56-71) | 67 | 41 (35-55) | 1 | 21 |
| Matched controls | 133 | 65 (56-70) | 67 | - | 0 | 2 |
| IPD > 4 years | 98 | 65 (56-73) | 66 | 67 (59-83) | 0 | 21 |
| Matched controls | 98 | 65 (56-72) | 66 | - | 0 | 3 |
| *LRRK2* nPD | 115 | 60 (56-65) | 49 | - | 0 | 3 |
| Matched controls | 115 | 60 (56-65) | 49 | - | 0 | 0 |
| *LRRK2* nPD < average age of onset | 52 | 56 (53-57) | 58 | - | 0 | 0 |
| Matched controls | 52 | 56 (53-57) | 58 | - | 0 | 0 |
| *LRRK2* nPD > average age of onset | 63 | 64 (62-68) | 40 | - | 0 | 6 |
| Matched controls | 63 | 64 (60-69) | 40 | - | 0 | 0 |
| *LRRK2* PD | 95 | 65 (57-70) | 56 | 55 (37-82) | 0 | 7 |
| Matched IPD | 95 | 65 (57-70) | 56 | 35 (23-59) | 1 | 18 |
| *LRRK2* PD | 95 | 65 (57-70) | 56 | 55 (37-82) | 0 | 7 |
| Matched *LRRK2* nPD | 95 | 65 (57-70) | 56 | - | 0 | 7 |
| *GBA* nPD | 109 | 63 (57-67) | 46 | - | 0 | 2 |
| Matched controls | 109 | 63 (57-67) | 46 | - | 0 | 1 |
| GC *GBA* nPD | 101 | 63 (57-67) | 47 | - | 0 | 2 |
| Matched controls | 101 | 60 (55-65) | 47 | - | 0 | 0 |
| *GBA* nPD > average age of onset | 91 | 64 (61-69) | 45 | - | 0 | 2 |
| Matched controls | 91 | 62 (57-66) | 45 | - | 0 | 1 |
| *GBA* PD | 127 | 62 (55-71) | 58 | 41 (23-65) | 1 | 18 |
| Matched IPD | 127 | 62 (55-71) | 58 | 35 (24-62) | 1 | 15 |
| *GBA* PD | 109 | 63 (57-67) | 54 | 58 (26-77) | 0 | 12 |
| Matched *GBA* nPD | 109 | 63 (57-67) | 54 | - | 0 | 2 |

In idiopathic Parkinson’s disease subjects who had been diagnosed more than 4 years previously, relatively high accuracies (88%, 95% CI [81%, 94%]) and AUC scores (0.89, 95% CI [0.83, 0.96]) were achieved. These scores reduced successively as duration from diagnosis decreased (Table 2), with scans undertaken less than one year from diagnosis yielding an accuracy of 67%, 95% CI [60%, 75%], and an AUC of 0.73, 95% CI [0.63, 0.83] (Supplementary Fig. 2). All idiopathic Parkinson’s disease models demonstrated similar regions of interest, notably in cerebrospinal fluid voxels surrounding the brainstem (Figure 1).

**Figure 1:**
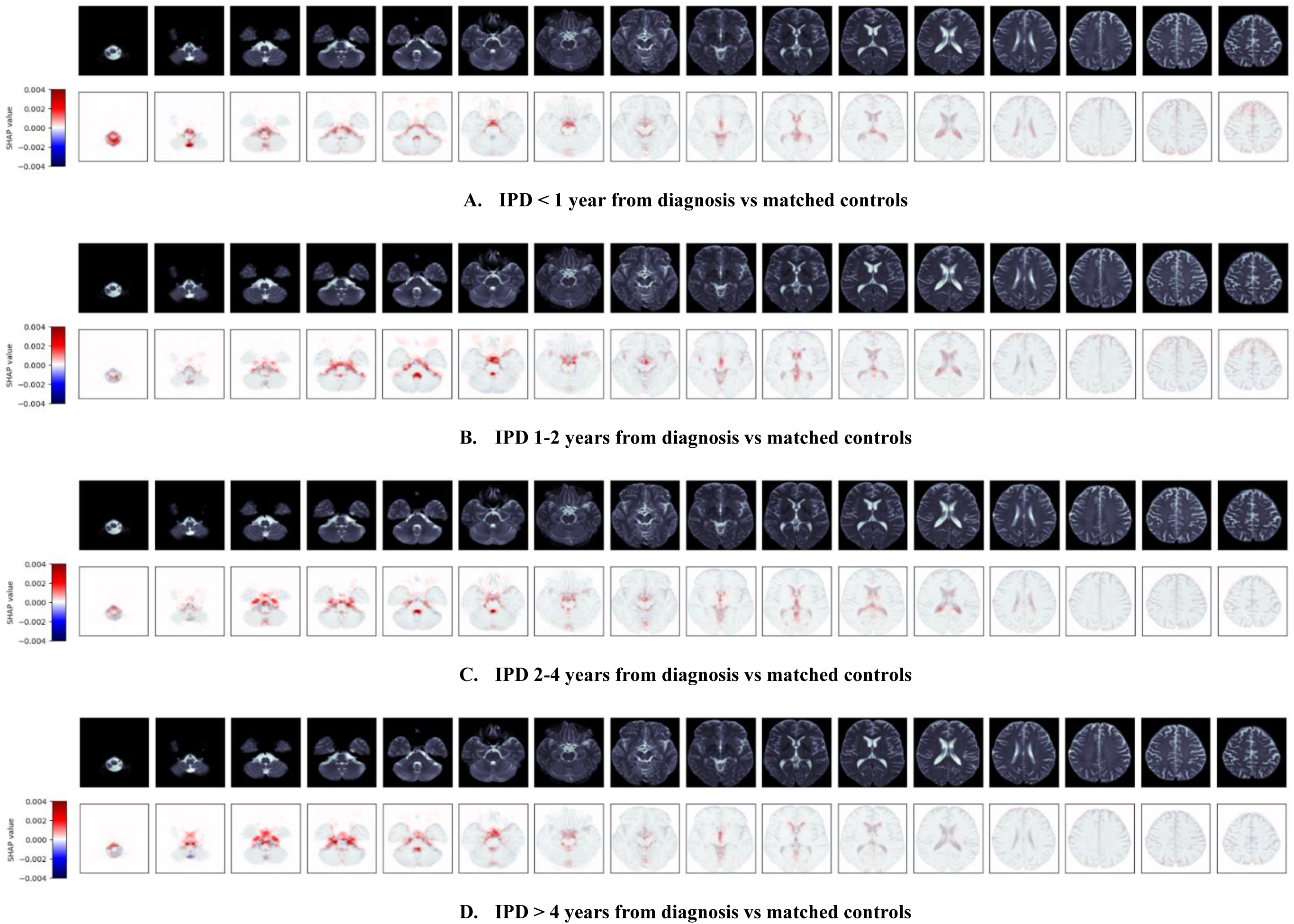
Mean SHapley Additive exPlanation (SHAP) maps for correct predictions of idiopathic Parkinson’s disease (IPD). Pixels highlighted in red have contributed to the prediction.

**Table 2:**
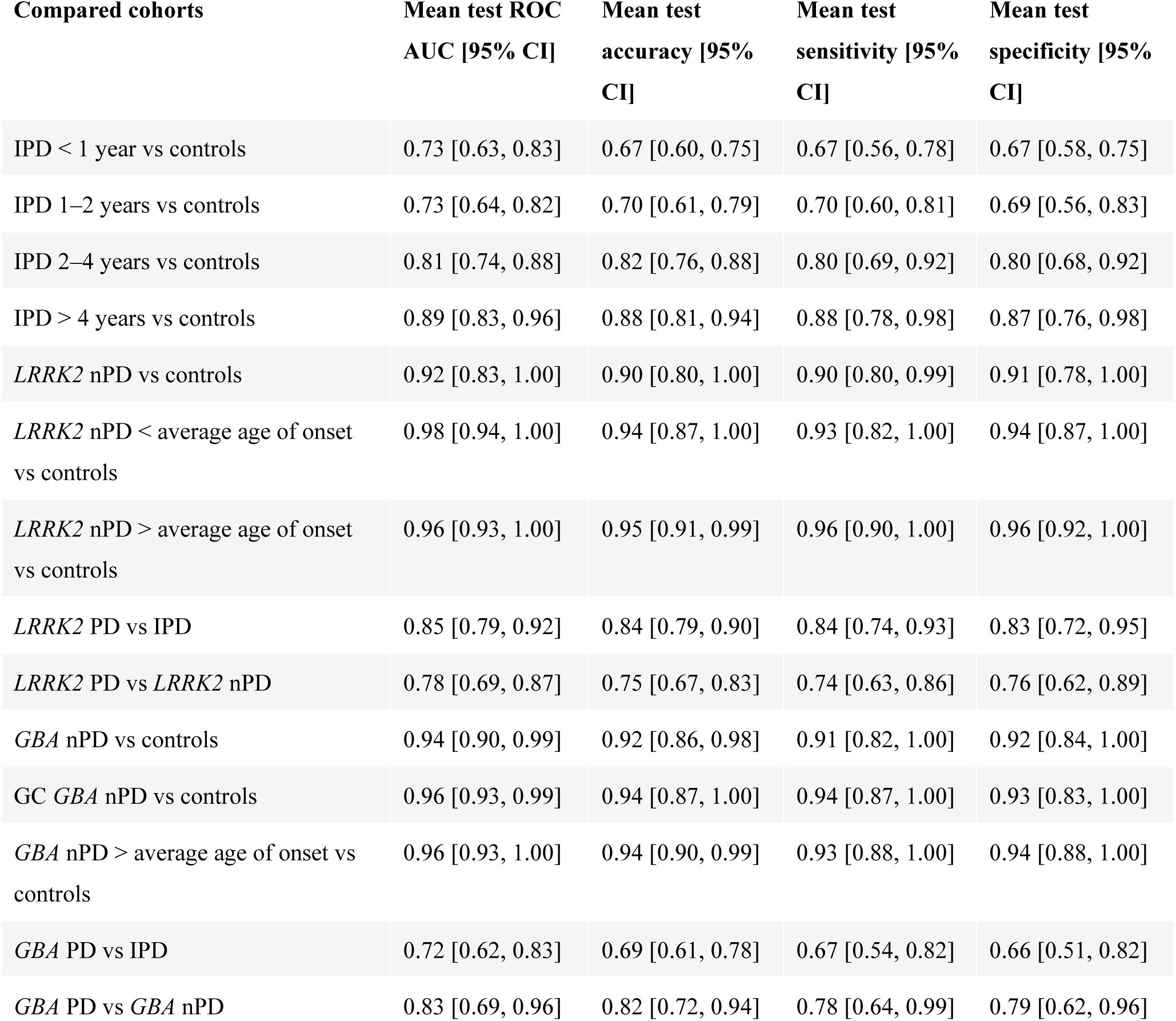
Model performance results. PD = Parkinson’s disease. IPD = idiopathic PD. *LRRK2* PD = *LRRK2* PD manifesting carriers. *LRRK2* nPD = *LRRK2* non PD manifesting carriers. *GBA* PD = *GBA* PD manifesting carriers. *GBA* nPD = *GBA* non PD manifesting carriers. GC *GBA* nPD = Gaucher causing *GBA* variants non manifesting carriers.

### LRRK2

In this analysis, 210 scans from 159 carriers of *LRRK2* risk variants were used. These were stratified into Parkinson’s disease manifesting carriers (*n*=95) and non-Parkinson’s disease manifesting carriers (*n*=115). In the case of the non-manifesting carriers, scans were further stratified by time of scan and divided into those acquired after the age of 59.4 (the average age of onset of *LRRK2* Parkinson’s disease)^9^ (*n*=63), and those taken before (*n*=52). Each pair of compared cohorts was matched on age and sex in a ratio of 1:1. Demographic data for these cohorts are shown in Table 1. Classification thresholds were chosen to maximise accuracy and balance sensitivity and specificity (Supplementary Fig. 6).

Models performed well in all cases (Table 2 and Supplementary Fig. 3). Ninety percent of non-manifesting *LRRK2*/control scans, 95% CI [80%, 100%], (AUC 0.92, 95% CI [0.83, 1.00]) were predicted correctly. This rose to 95%, 95% CI [91%, 99%] (AUC 0.96, 95% CI [0.95, 1.00]) in the scans from non-manifesting *LRRK2* subjects over the age of onset. Notably the model comparing *LRRK2* and idiopathic Parkinson’s disease performed well, predicting *LRRK2* scans with 84% accuracy, 95% CI [79%, 90%] (AUC 0.85, 95% CI [0.79, 0.92]). Average SHAP heatmaps demonstrated predominant interest in pixels immediately adjacent to the brainstem parenchyma (Figure 2).

**Figure 2.**
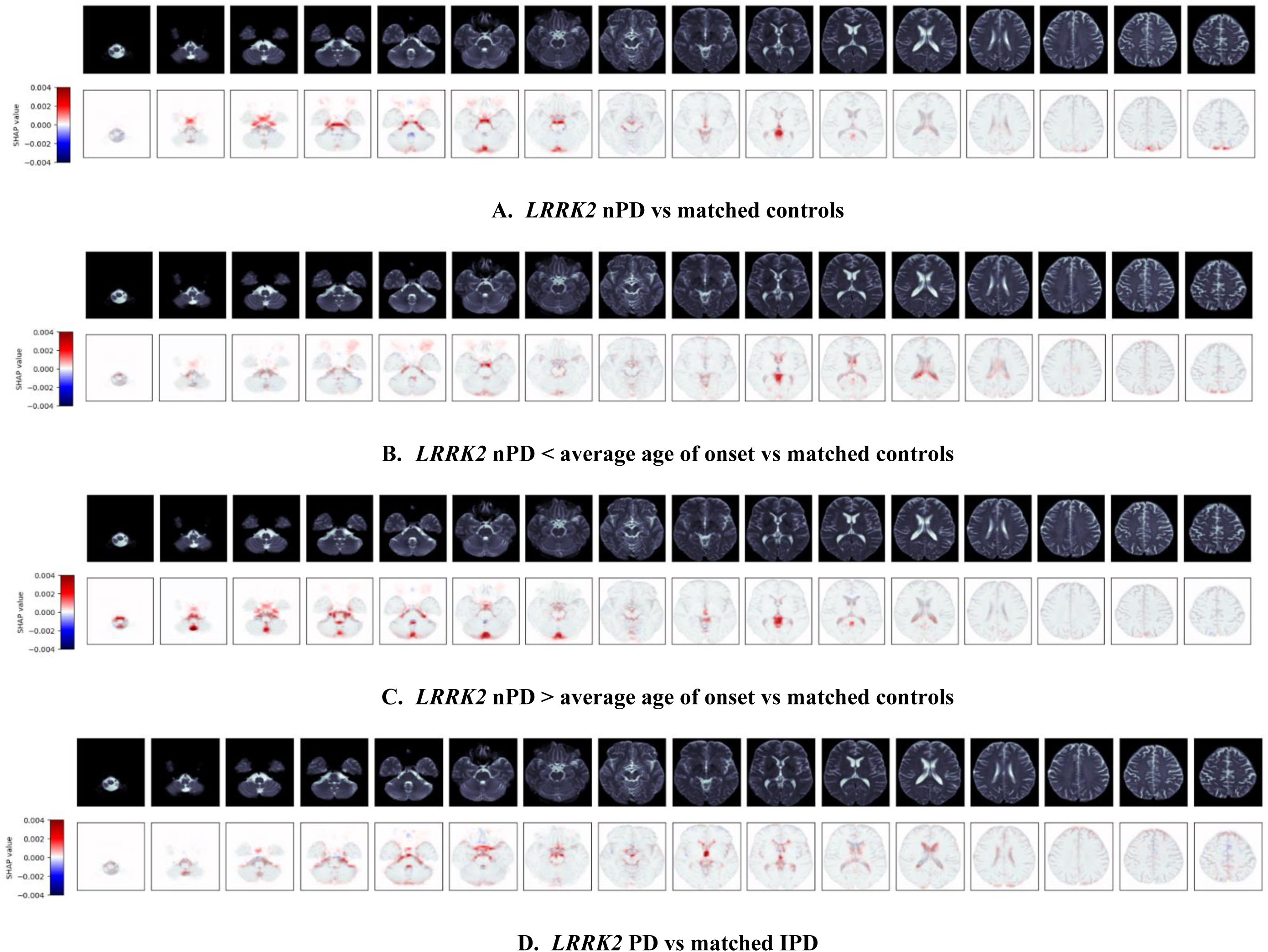
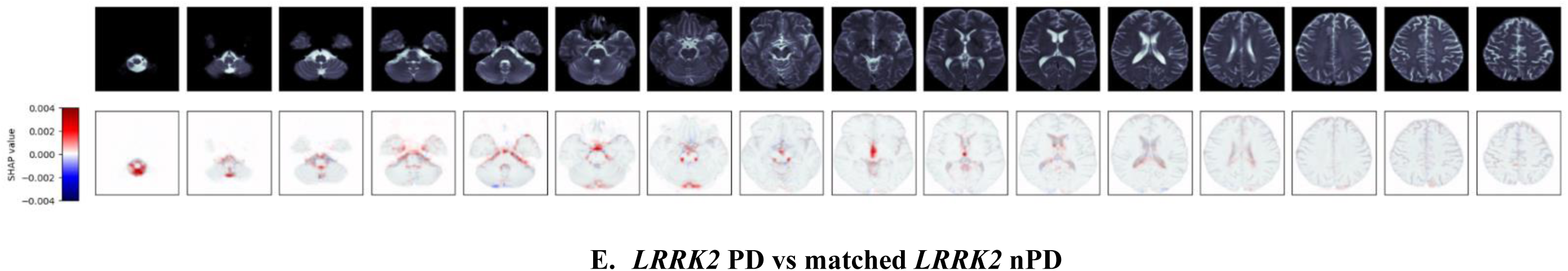
Mean SHapley Additive exPlanation (SHAP) maps for correct predictions of *LRRK2* carriers. Pixels highlighted in red have contributed to the prediction. *LRRK2* PD = *LRRK2* Parkinson’s disease manifesting carriers. *LRRK2* nPD = *LRRK2* non Parkinson’s disease manifesting carriers. IPD = idiopathic Parkinson’s disease.

### GBA

In this analysis, 236 scans from 159 carriers of a *GBA* variant were used. These were stratified into manifesting carriers (*n*=127) and non-manifesting carriers (*n*=109). The scans were also stratified into 184 scans from carriers of Gaucher causing *GBA* variants and 51 scans from carriers of the non-Gaucher causing variants p.E326K (alternative nomenclature p.E365K) and p.T369M (p.T409M). There were insufficient numbers to build models using cases stratified into ‘mild’ and ‘severe’ variants. The non-manifesting *GBA* group was also further stratified by time of scan: divided into those acquired after the age of 55.8 (the average age of onset of *GBA* related Parkinson’s disease)^9^ (*n*=91), and those taken before (*n*=18). The latter cohort was not large enough to build a model. Each pair of compared cohorts was matched on age and sex in a ratio of 1:1. Demographic data for these cohorts are shown in Table 1. Classification thresholds were chosen to maximise accuracy and balance sensitivity and specificity (Supplementary Fig. 7)

*GBA* models performed well (Table 2 and Supplementary Fig. 4). The model built for the combined non-manifesting *GBA* cohort was able to successfully predict *GBA* scans with an accuracy of 92%, 95% CI [86%, 98%] (AUC 0.94, 95% CI [0.90, 0.99]). This increased marginally to 94%, 95% CI [87%, 100%] (AUC 0.96, 95% CI [0.93, 0.99]) in the combined ‘mild’ and ‘severe’ non-manifesting *GBA* cohort. In the cohort of non-manifesting *GBA* participants over the average age of onset, performance was slightly higher than the main model (94% accuracy, 95% CI [90%, 99%], AUC 0.96, 95% CI [0.93, 1.00]). In common with idiopathic Parkinson’s, SHAP heatmaps demonstrated interest in non-parenchymal pixels surrounding the brainstem (Figure 3). Additionally, there was a focus on pixels adjacent to the posterior occipital lobe.

**Figure 3.**
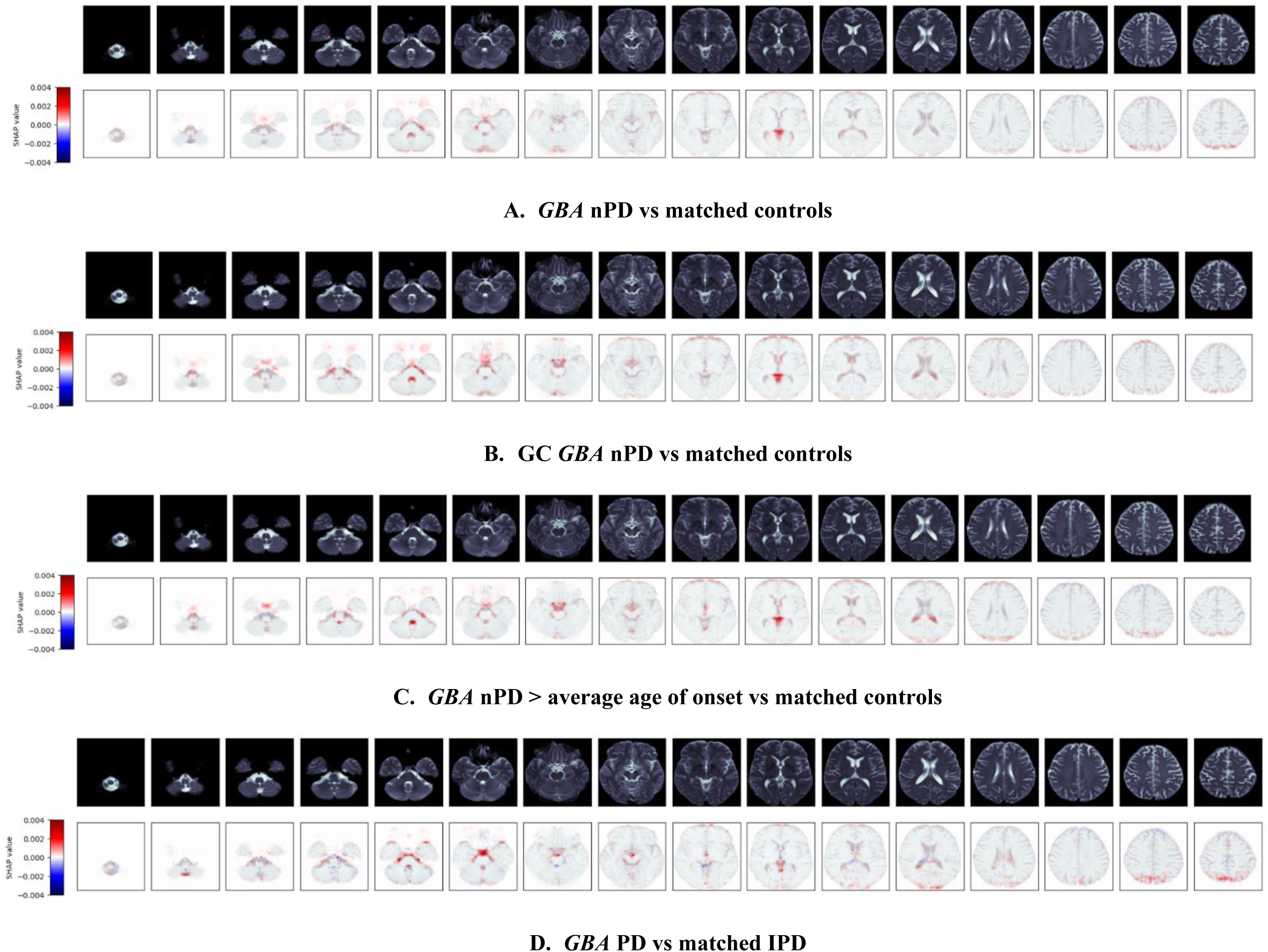
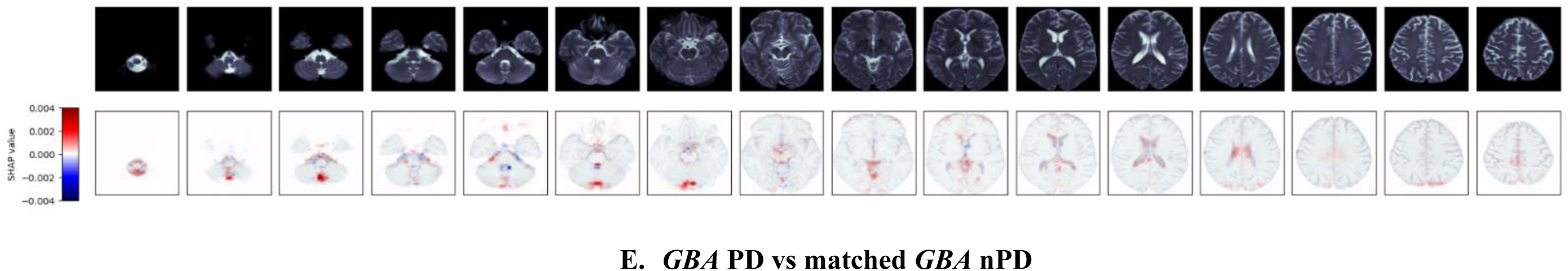
Mean SHapley Additive exPlanation (SHAP) maps for correct predictions of *GBA* carriers. Pixels highlighted in red have contributed to the prediction. *GBA* PD = *GBA* Parkinson’s disease manifesting carriers. *GBA* nPD = *GBA* non Parkinson’s disease manifesting carriers. GC *GBA* nPD = Gaucher causing *GBA* variants non manifesting carriers. IPD = idiopathic Parkinson’s disease.

## Discussion

Our models reliably differentiated Parkinson’s disease from control scans. The findings suggest that deep learning can identify progressive changes in both idiopathic and genetic Parkinson’s disease. The trained models exhibited good performance for established cases of idiopathic Parkinson’s disease, which steadily decreased for models trained on earlier cases of Parkinson’s disease.

Most of the interest of our models appears focused on cerebrospinal fluid spaces surrounding the brainstem. Models might have detected enlargement in the cerebrospinal fluid spaces caused by cortical atrophy. Cerebral atrophy has previously been found in Parkinson’s disease, with reported reductions in grey matter volume in the frontal and temporal lobes,^23^ and diffuse gyral atrophy throughout the temporal, parietal and frontal cortices.^24^ A recent study has found enlargement of the interpeduncular and right ambient cisterns in patients with Parkinson’s disease.^25^ Another possibility is that the models have detected ventricular enlargement caused by cortical changes. Asymmetric lateral ventricular enlargement has been reported in Parkinson’s disease, associated with progression.^26^ Ventricular enlargement has also been associated with cognitive decline in Parkinson’s disease.^27^ Such progressive patterns might explain the models’ higher performance for later stages of Parkinson’s disease.

To investigate whether these changes were visible in the genetic cohorts, we built models for the non-manifesting carriers of *LRRK2* and *GBA* variants, both genetic risk factors for Parkinson’s disease. Between 28–74% of *LRRK2* and 8–10% of *GBA* carriers will develop Parkinson’s disease,^7,8^ hence a significant portion of these subjects would be expected to be within the prodromal phase. Given the findings of our idiopathic Parkinson’s disease models, we predicted that these models would not perform well, as any brain changes were anticipated to be early and subtle. However, the performance of these models was remarkably high for both non-manifesting *GBA* and non-manifesting *LRRK2* groups. Once again, explainability techniques suggested a focus on pixels adjacent to the brainstem. In the case of the non-manifesting *GBA* carriers, the models also demonstrated interest in pixels adjacent to the posterior parieto-occipital lobe. *GBA*-associated Parkinson’s disease has been shown to be associated with a higher frequency of cognitive deficits in the visuospatial domain as well as visual hallucinations,^28^ hence atrophic changes within the wider visual processing regions would seem to be of particular relevance.

We are unable to make more certain judgement on the drivers of these very high model performances for both non-manifesting *GBA* and non-manifesting *LRRK2*. The high performance does however suggest that there is scope to use such techniques to identify carriers of these genetic risk factors using machine learning models. It may also suggest that early brain changes in *GBA* and *LRRK2* carriers are distinct from early brain changes in idiopathic Parkinson’s disease, which would support speculation that genetic and idiopathic Parkinson’s disease are separate disease pathways that converge on a broadly shared phenotype.^29^

To further investigate these findings for potential progressive changes, we subdivided the non-manifesting *GBA* and *LRRK2* cohorts again by age, to assess whether there was any difference in the performance of models built to distinguish scans before and after the average age of onset of *GBA*/*LRRK2* Parkinson’s disease. Unfortunately, among the *GBA* carriers, there were only enough scans taken after the average age of onset to build models for these. These had a slightly higher performance than models built for all *GBA* non-manifesting ages. For the *LRRK2* carriers, models built for scans from the older carriers yielded a marginally higher accuracy than the younger carriers. In both cases this may suggest that the brain differences become more pronounced with time.

### Limitations

The size of the dataset is a limitation of this research. Dividing the PPMI dataset according to scan type, genetic status, and timing of scan in relation to diagnosis produces subgroups that may be too small to reliably train classification models. If it were possible to obtain more data, this might enable the development of even more accurate and generalisable models. We have mitigated this limitation to an extent by augmenting the training data with horizontal flip, thus artificially increasing the size of the dataset using a well-established technique.^30^

A further limitation of this study is the likelihood of variability in the idiopathic Parkinson’s disease cohort. Imaging from these subjects will be more heterogeneous in terms of disease cause and MRI profile than imaging from the genetic Parkinson’s disease cohorts. This may partially explain the comparatively high performances for the models trained on the genetic Parkinson’s imaging.

Another limitation is the lack of external validation. External validation sets are difficult to obtain as appropriate publicly available databases do not exist. Our research team is in the process of planning and gaining governance clearance for such accessible studies. In this study we have mitigated this limitation as far as possible by using ten-fold cross-validation and reporting the mean validation metrics. However, the validation data originate from the same source as the training data, and the metrics reported may not be representative of the models’ performance on data from a different distribution. For example, these models trained on a controlled research dataset may not generalise well to data routinely collected in a clinical setting. An external validation set would allow for more accurate assessment of the models’ capability to generalise to other populations.

### Future work

In the future, these results should be validated in an external dataset. Our research team is in the process of collecting such a dataset of routinely-collected brain imaging. Routinely-collected imaging from carriers of genetic variants associated with Parkinson’s disease would be more difficult to collect, but would also be of value to validate these results.

In addition, the inclusion of more data sources might improve predictive performance. Other modalities and functional imaging should be considered in future work.

## Conclusions

The findings suggest that deep learning can identify progressive changes in both idiopathic and genetic Parkinson’s disease. Differences in the brains of non-manifesting genetic variant carriers are even more obvious, which may reflect distinct starting points and progression pathways in genetic Parkinson’s disease.

## Data availability

Data used in the preparation of this article were obtained from the Parkinson’s Progression Markers Initiative (PPMI) database (https://www.ppmi-info.org/access-data-specimens/download-data). For up-to-date information on the study, visit https://www.ppmi-info.org. All data used in this study, as well as a data dictionary, are free and publicly available at the PPMI website. Additional related documents including the study protocol and assay methods are also available. Data access can be requested on the website. There are no restrictions on who can request access.

## Notes

**Conflicts of interest:** The authors report no conflicts of interest.

### Competing Interest Statement

The authors have declared no competing interest.

### Author Declarations

The study used only openly available human data from the Parkinson's Progression Markers Initiative.

