## Supplementary Material for "Deep learning classification of MRI differentiates brain changes in genetic and idiopathic Parkinson’s disease"

| *LRRK2* | |
|---|---|
| **Variant** | **Participant count** |
| p.S1647T/p.S1647T / p.G2019S / p.M2397T/p.M2397T | 37 |
| p.S1647T / p.G2019S / p.M2397T | 31 |
| p.S1647T / p.G2019S / p.M2397T/p.M2397T | 20 |
| p.I723V / p.S1647T / p.G2019S / p.M2397T | 12 |
| p.N551K / p.R1398H / p.S1647T / p.G2019S / p.M2397T/p.M2397T | 11 |
| p.G2019S | 11 |
| p.S1647T / p.G2019S / p.N2081D / p.M2397T | 6 |
| p.N551K / p.S1647T/p.S1647T / p.G2019S / p.M2397T/p.M2397T | 4 |
| p.P1542S / p.S1647T / p.G2019S / p.M2397T/p.M2397T | 3 |
| p.R1441G / p.M1646T / p.M2397T | 3 |
| p.M1646T / p.S1647T / p.G2019S / p.M2397T/p.M2397T | 2 |
| p.N551K / p.R1398H / p.R1441G / p.M1646T / p.M2397T/p.M2397T | 2 |
| p.S1647T/p.S1647T / p.G2019S/p.G2019S / p.M2397T/p.M2397T | 2 |
| p.I723V / p.S1647T / p.G2019S / p.M2397T/p.M2397T | 2 |
| p.I723V / p.M1646T / p.S1647T / p.G2019S / p.M2397T/p.M2397T | 2 |
| p.R1441G | 2 |
| p.R1441G / p.M1646T / p.S1647T / p.M2397T/p.M2397T | 2 |
| p.R1514Q / p.S1647T / p.G2019S / p.M2397T/p.M2397T | 2 |
| p.N59K / p.S1647T / p.G2019S / p.M2397T | 1 |
| p.N551K / p.S1647T / p.G2019S / p.M2397T | 1 |
| p.N551K / p.R1398H / p.R1441C / p.S1647T / p.M2397T/p.M2397T | 1 |
| p.I723V / p.S1647T/p.S1647T / p.G2019S / p.M2397T/p.M2397T | 1 |
| p.V1330M / p.S1647T / p.G2019S / p.M2397T | 1 |
| ***GBA*** | |
| **PD risk factor *GBA* variant** | |
| **Variant** | **Participant count** |
| p.E326K (p.E365K) | 11 |
| p.T369M (p.T408M) | 8 |
| p.T336S | 1 |
| **'Mild' Gaucher causing *GBA* variant** | |
| **Variant** | **Participant count** |
| p.N370S (p.N409S) | 113 |

| | |
|---|---|
| p.N370S/p.N370S (p.N409S/p.N409S) | 8 |
| p.K13R | 4 |
| p.R44C (p.R83C) | 1 |
| p.A456P (p.A495P) | 1 |
| p.R39C (p.R78C) | 1 |
| p.E365K/p.N370S (p.E365K/p.N409S) | 1 |
| p.G115R/p.G193E (p.G154R/p.G232E) | 1 |
| p.I489L (p.I528L) | 1 |
| **'Severe' Gaucher causing *GBA* variant** | |
| **Variant** | **Participant count** |
| p.L444P (p.483P) | 5 |
| p.IVS2+1G>A | 1 |
| p.T369M/p.R120W (p.T408M/p.R159W) | 1 |
| p.R502C | 1 |

**Supplementary Table 1: *LRRK2* and *GBA* variants present in cohorts**

| | Participant count | | | |
|---|---|---|---|---|
| | **PD** | **Control** | ***LRRK2*** | ***GBA*** |
| Total included in analysis | 193 | 193 | 159 | 159 |
| **Withdrawal reason** | | | | |
| Subject withdrew consent | 29 | 18 | 10 | 15 |
| Death | 15 | 2 | 3 | 10 |
| Lost to follow up | 12 | 13 | 4 | 2 |
| Other | 10 | 3 | 5 | 8 |
| Informant / Caregiver decision | 3 | 2 | 2 | 4 |
| Investigator decision | 3 | 1 | 0 | 0 |
| Decline in health | 2 | 1 | 0 | 0 |
| Transportation/Travel issues (ex: logistics or travel, moved away from study site) | 2 | 0 | 0 | 1 |
| Family, care-partner, or social issues (such as work/job obligations) | 1 | 0 | 2 | 0 |
| Adverse Event | 1 | 0 | 0 | 0 |
| Burden of study procedures (other than travel) | 0 | 1 | 0 | 0 |
| Sponsor decision | 0 | 0 | 1 | 1 |
| Institutionalised | 0 | 0 | 0 | 1 |
| **Total withdrawals** | 78 | 41 | 27 | 42 |
| | 188 | | | |

**Supplementary Table 2: Counts of participants included in this analysis and reasons for withdrawal**

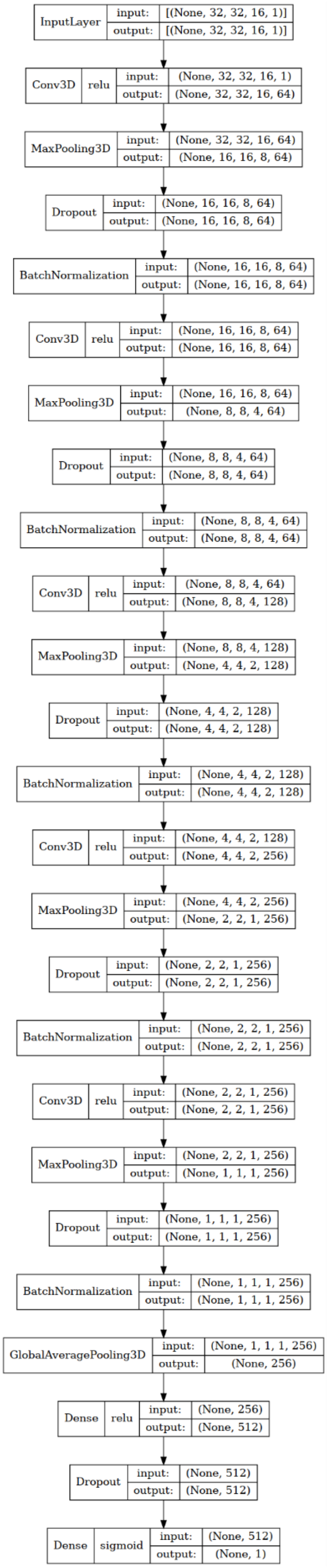


**Supplementary Figure 1: Convolutional neural network architecture**

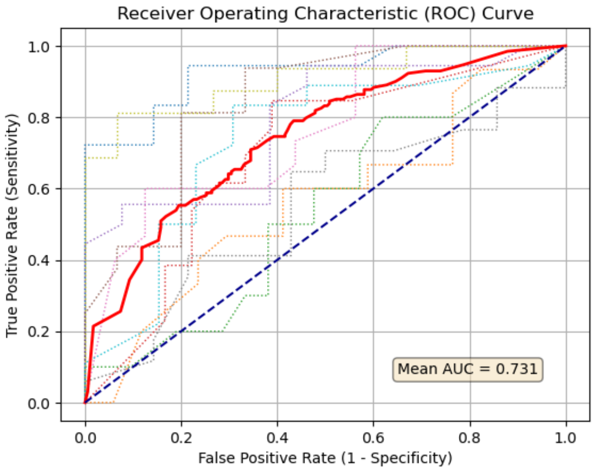


A. **IPD < 1 year vs matched controls**

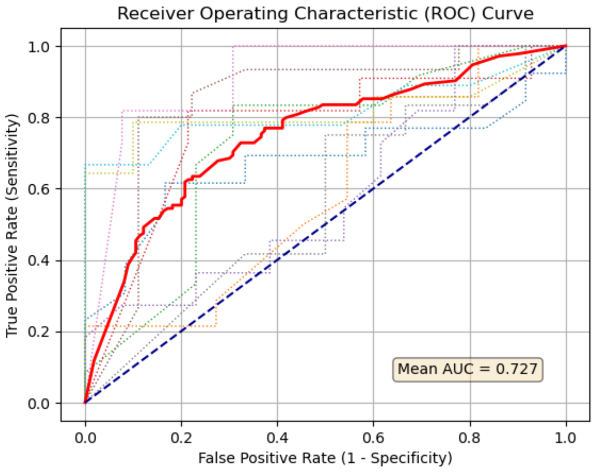


B. **IPD 1-2 years vs matched controls**

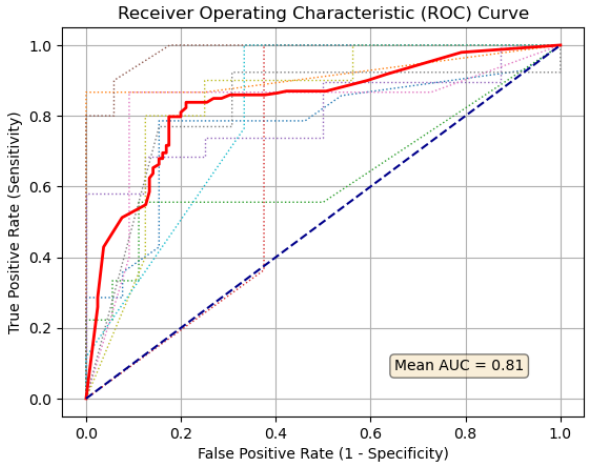


C. **IPD 2-4 years vs matched controls**

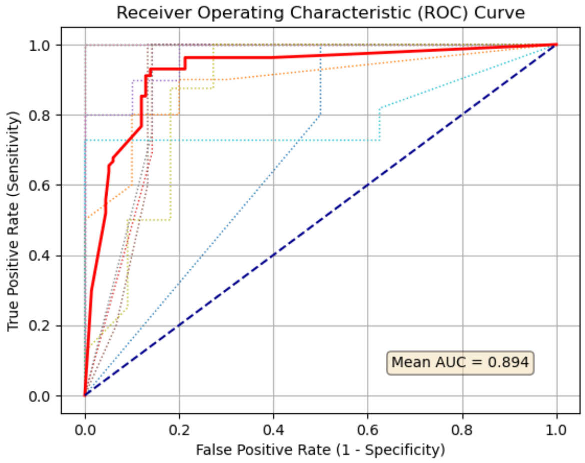


D. **IPD > 4 years vs matched controls**

**Supplementary Figure 2: Receiver Operating Characteristic (ROC) curves for idiopathic Parkinson's disease (IPD) models.** The bold red line represents the mean ROC curve; the dotted lines represent the ROC curve per k-fold.

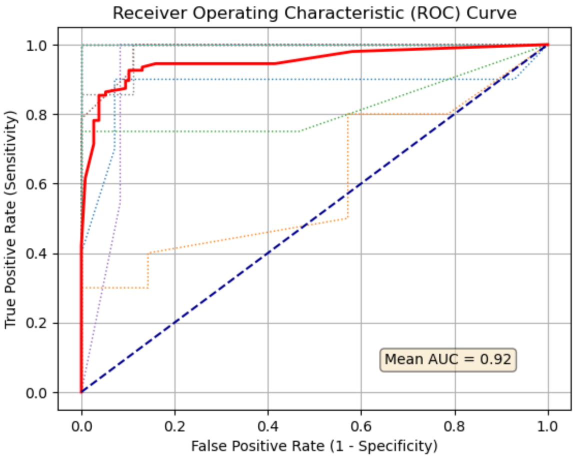


A. ***LRRK2*** **nPD vs matched controls**

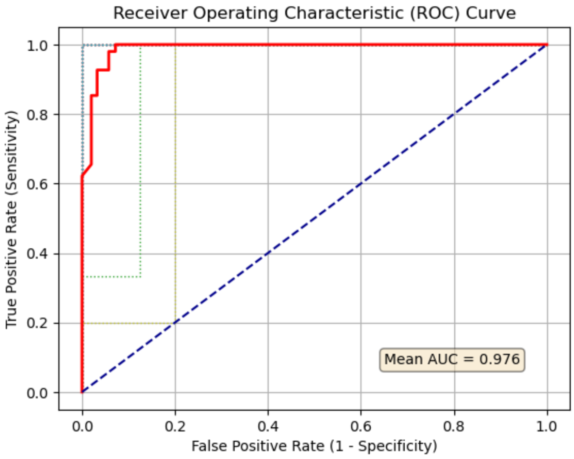


B. ***LRRK2*** **nPD < average age of onset vs matched controls**

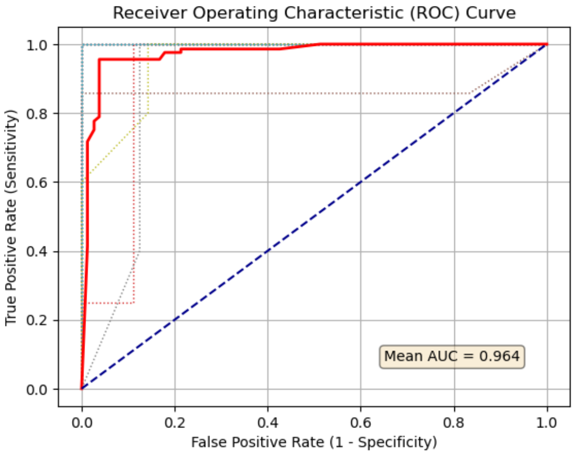


C. ***LRRK2*** **nPD > average age of onset vs matched controls**

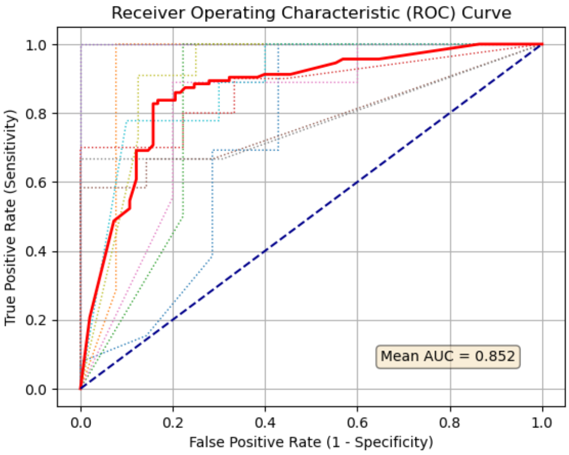


D. ***LRRK2*** **PD vs matched IPD**

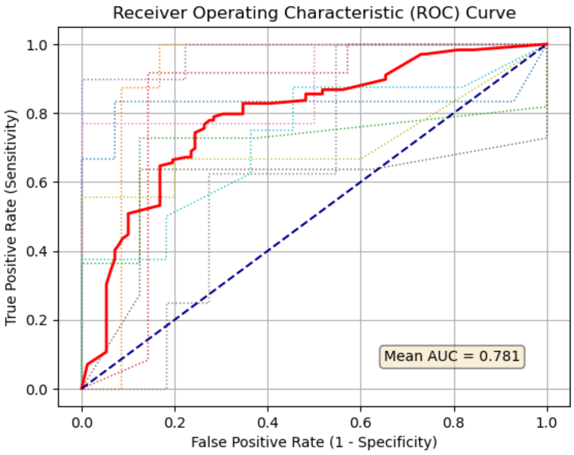


E. ***LRRK2*** **PD vs matched** ***LRRK2*** **nPD**

**Supplementary Figure 3: Receiver Operating Characteristic (ROC) curves for *LRRK2* models.** The bold red line represents the mean ROC curve; the dotted lines represent the ROC curve per k-fold. *LRRK2* PD = *LRRK2* Parkinson's disease manifesting carriers. *LRRK2* nPD = *LRRK2* non Parkinson's disease manifesting carriers. IPD = idiopathic Parkinson's disease.

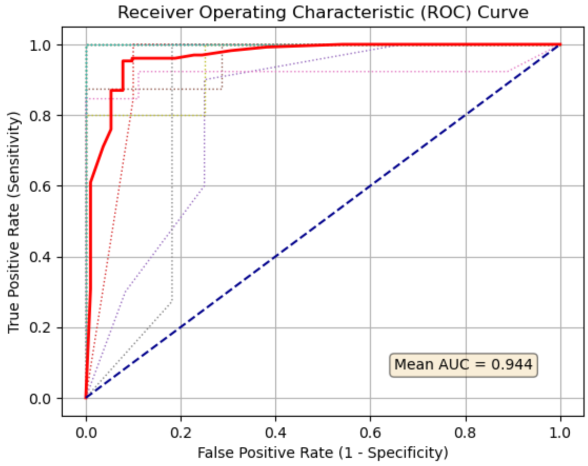


A. ***GBA* nPD vs matched controls**

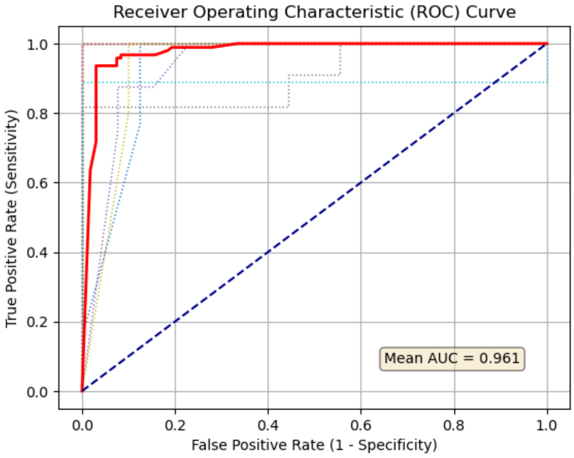


B. **GC *GBA* nPD vs matched controls**

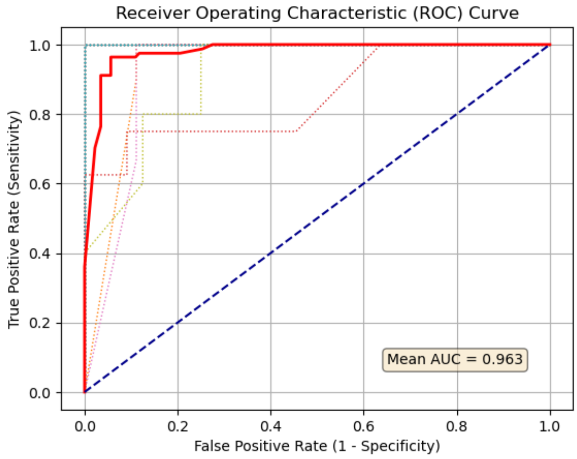


C. ***GBA* nPD > average age of onset vs matched controls**

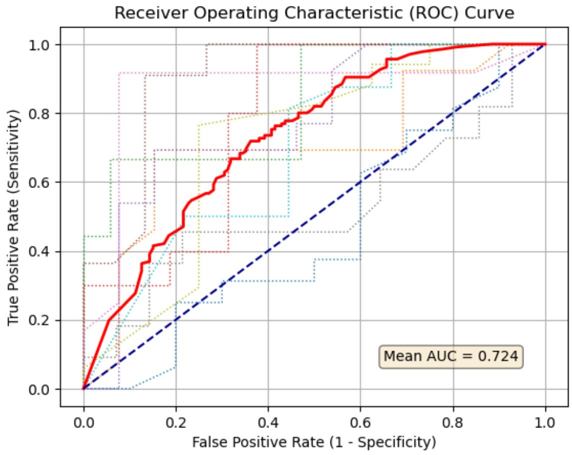


D. ***GBA* PD vs matched IPD**

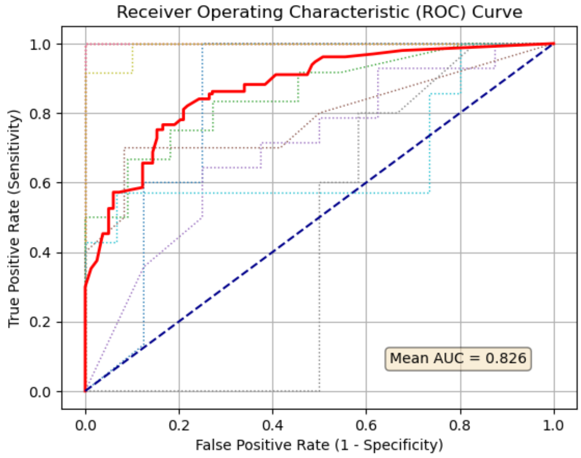


E. ***GBA* PD vs matched *GBA* nPD**

**Supplementyar Figure 4: Receiver Operating Characteristic (ROC) curves for *GBA* models.** The bold red line represents the mean ROC curve; the dotted lines represent the ROC curve per k-fold. *GBA* PD = *GBA* Parkinson's disease manifesting carriers. *GBA* nPD = *GBA* non Parkinson's disease manifesting carriers. GC *GBA* nPD = Gaucher causing *GBA* variants non manifesting carriers. IPD = idiopathic Parkinson's disease.

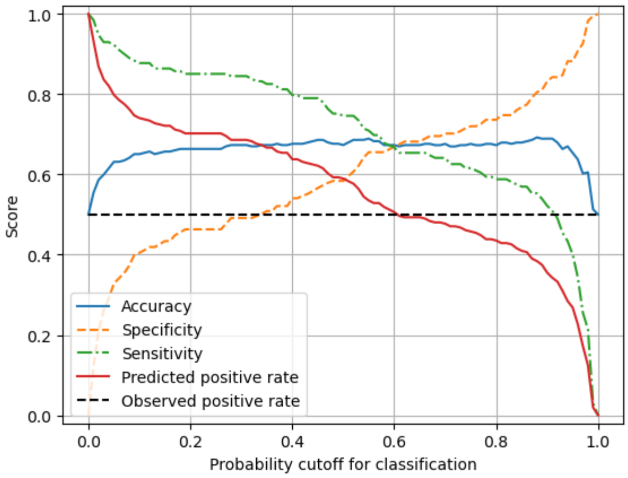


**A. IPD < 1 year vs matched controls. Threshold chosen: 0.60.**

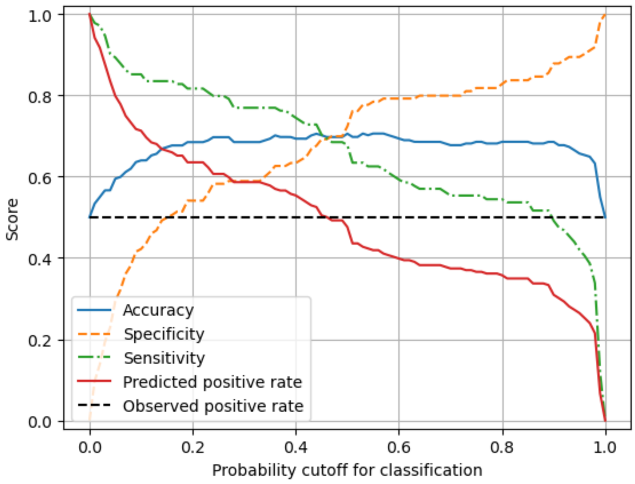


**B. IPD 1-2 years vs matched controls. Threshold chosen: 0.45.**

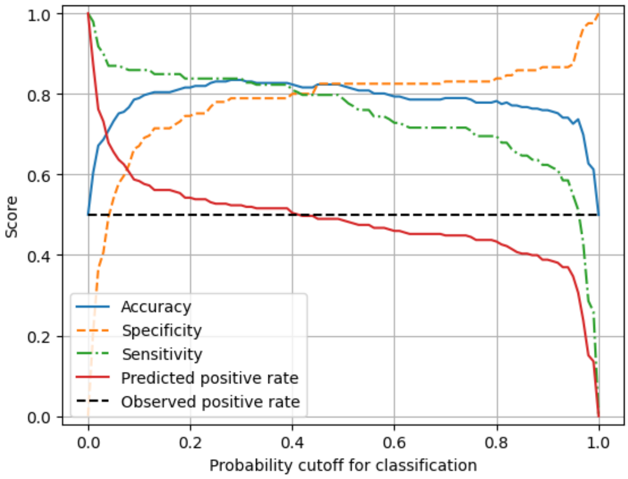


**C. IPD 2-4 years vs matched controls. Threshold chosen: 0.41.**

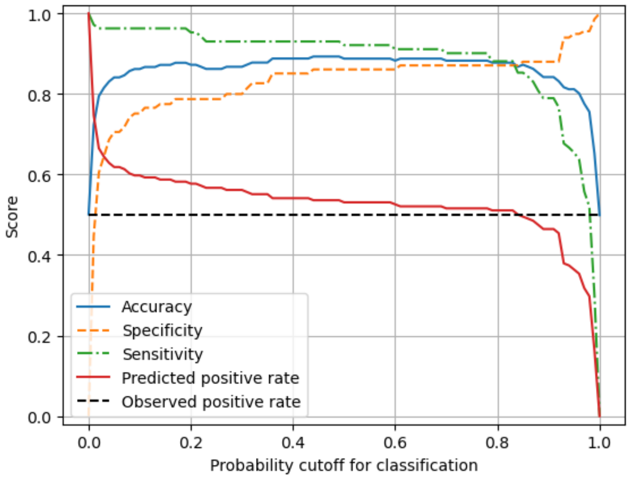


**D. IPD > 4 years vs matched controls. Threshold chosen: 0.83.**

**Supplementary Figure 5: Mean test performance metrics for idiopathic Parkinson's disease (IPD) models**

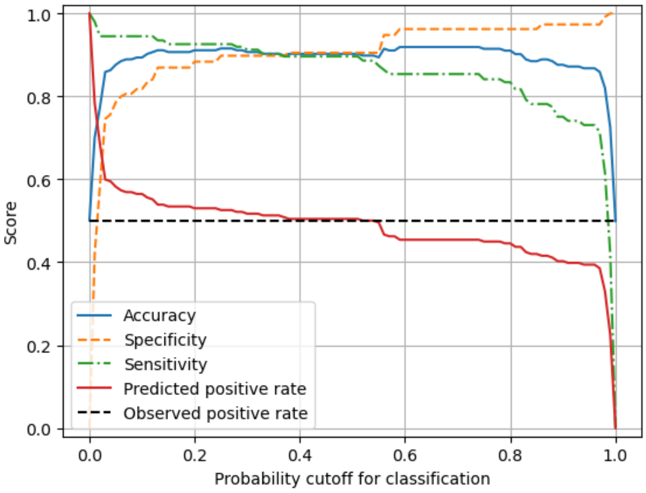


**A.** ***LRRK2*** **nPD vs matched controls. Threshold chosen: 0.38.**

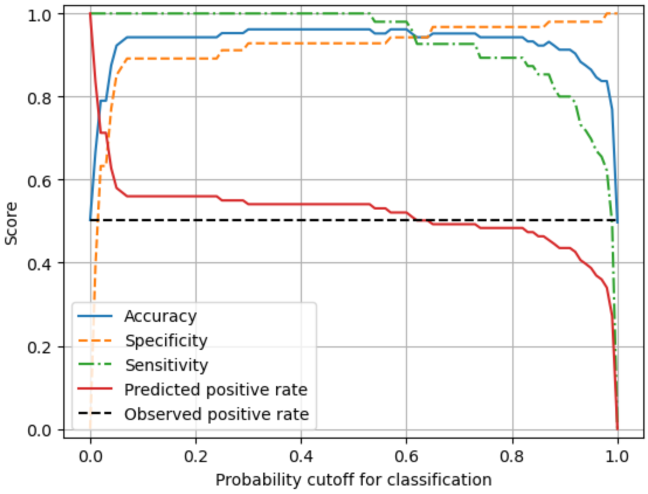


**B.** ***LRRK2*** **nPD < average age of onset vs matched controls. Threshold chosen: 0.62.**

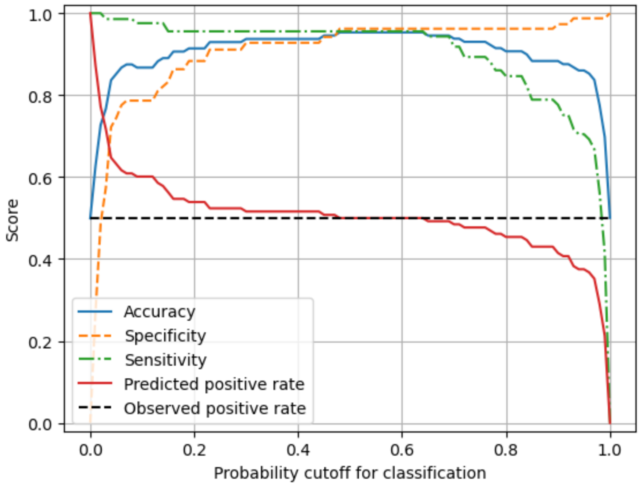


**C.** ***LRRK2*** **nPD > average age of onset vs matched controls. Threshold chosen: 0.5.**

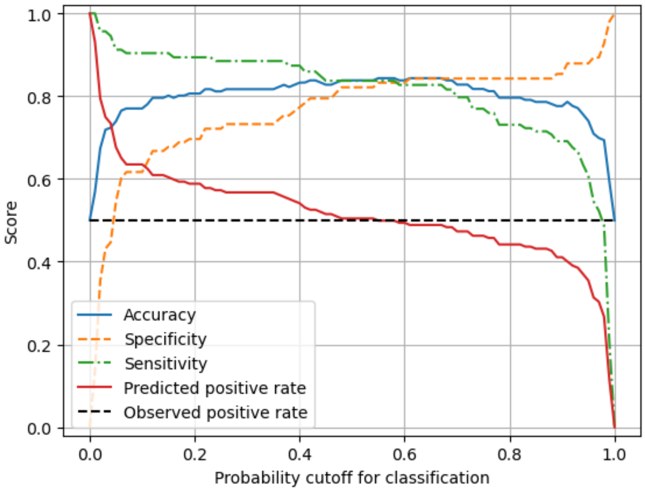


**D.** ***LRRK2*** **PD vs matched IPD. Threshold chosen: 0.55.**

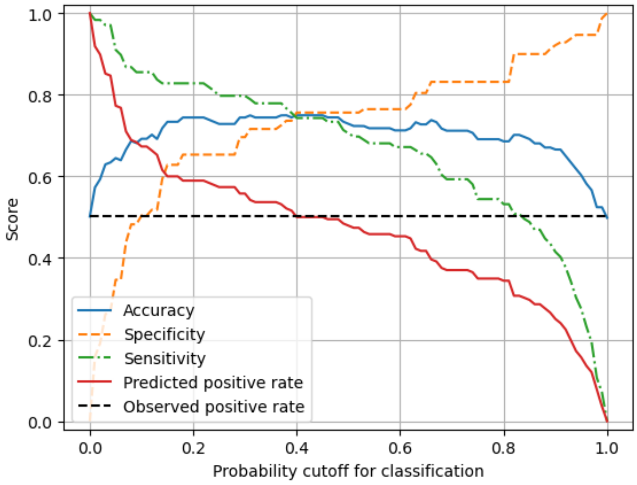


**E.** ***LRRK2*** **PD vs matched** ***LRRK2*** **nPD. Threshold chosen: 0.40.**

**Supplementary Figure 6: Mean test performance metrics for** ***LRRK2*** **models.** *LRRK2* PD = *LRRK2* Parkinson's disease manifesting carriers. *LRRK2* nPD = *LRRK2* non Parkinson's disease manifesting carriers. IPD = idiopathic Parkinson's disease.

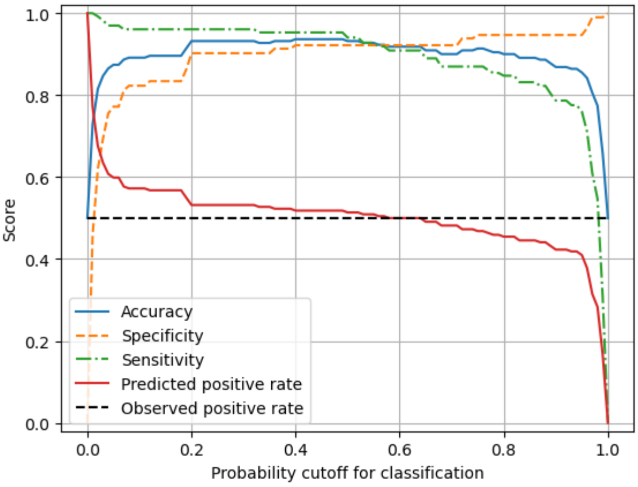


**A. *GBA* nPD vs matched controls. Threshold chosen: 0.58.**

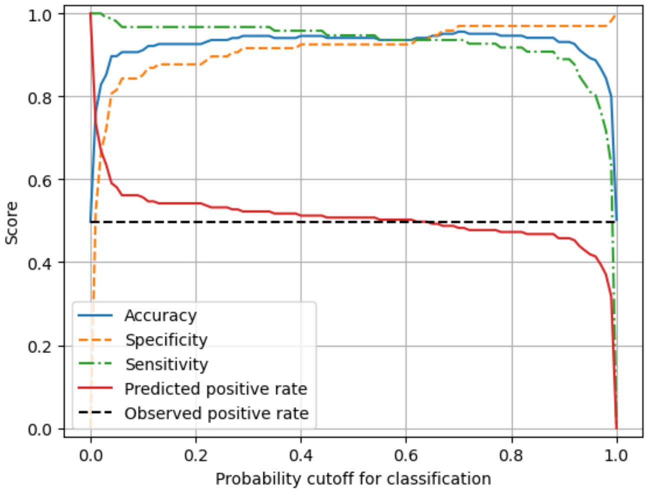


**B. GC *GBA* nPD vs matched controls. Threshold chosen: 0.60.**

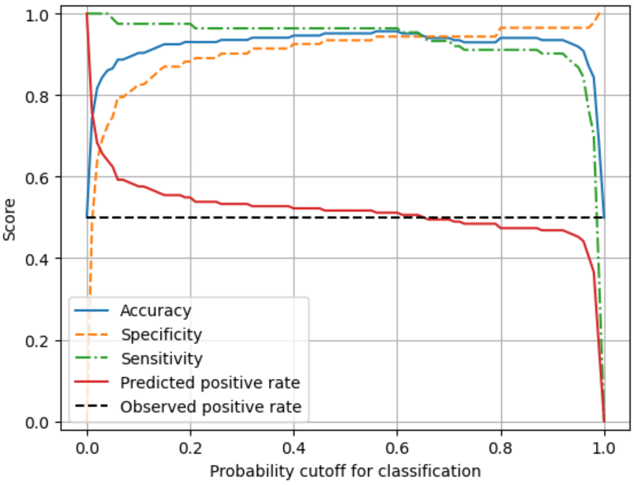


**C. *GBA* nPD > average age of onset vs matched controls. Threshold chosen: 0.65.**

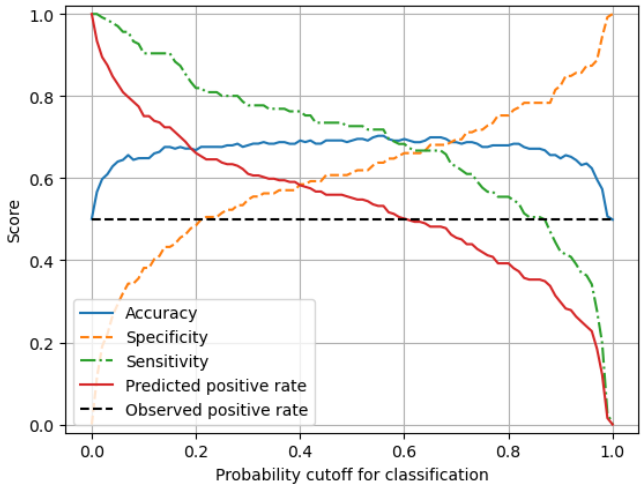


**D. *GBA* PD vs matched IPD. Threshold chosen: 0.60.**

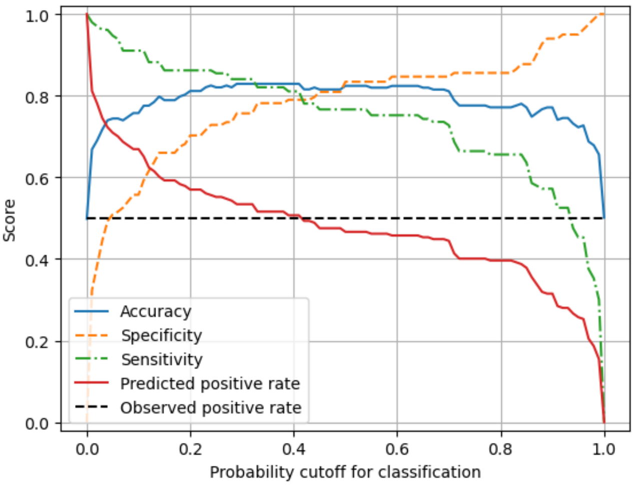


**E. *GBA* PD vs matched *GBA* nPD. Threshold chosen: 0.41.**

**Supplementary Figure 7: Mean test performance metrics for *GBA* models.** *GBA* PD = *GBA* Parkinson's disease manifesting carriers. *GBA* nPD = *GBA* non Parkinson's disease manifesting carriers. GC *GBA* nPD = Gaucher causing *GBA* variants non manifesting carriers. IPD = idiopathic Parkinson's disease.
